# Relationship between blood bone metabolic biomarkers and anemia in CKD patients

**DOI:** 10.1101/2022.12.13.22283190

**Authors:** Fan Li, Guang Yang, Xiaoxue Ye, Ming Zeng, Hui Huang, Anning Bian, Changying Xing, Shaowen Tang, Jing Zhang, Yao Jiang, Huimin Chen, Caixia Yin, Lina Zhang, Jing Wang, Yaoyu Huang, Wenbin Zhou, Huiting Wan, Xiaoming Zha, Ningning Wang

## Abstract

**Introduction:** Blood bone metabolic biomarkers are non-invasive indexes for evaluating renal osteodystrophy (ROD). Here the relationships between blood bone metabolic biomarkers and anemia in chronic kidney disease (CKD) patients are investigated, the effects of parathyroidectomy (PTX) on above indices are analyzed.

**Methods:** In this cross-sectional study, 100 healthy controls and 239 CKD patients, including 46 secondary hyperparathyroidism (SHPT) subgroup with PTX, were enrolled. A prospective study with 28 PTX patients was followed up. The degree of anemia was classified as mild, moderate, and severe based on the tertiles of hemoglobin (Hb) levels of the anemic CKD patients, with cutoff values of 83g/L and 102g/L. Bone metabolic biomarkers, including calcium (Ca), phosphorus (P), intact parathyroid hormone (iPTH), fibroblast growth factor 23 (FGF23) and α-klotho were tested.

**Results:** The mean eGFR in CKD patients was 25.74±35.99 ml/min/1.73 m^2^ and 84.10% patients had anemia. The baseline Hb levels in the mild, moderate, and severe anemia subgroups were 110.86±5.99g/L, 92.71±5.96g/L and 67.38±10.56g/L, respectively. CKD patients had higher adjusted Ca, P, ALP, iPTH and FGF23 levels, and lower α-klotho levels than controls. Baseline adjusted Ca, P, iPTH and α-klotho levels were associated with Hb in CKD patients. Blood adjusted Ca, P, iPTH levels were correlated with anemia severity. After PTX (median interval: 6.88 months), anemia and high blood adjusted Ca, P, iPTH and FGF23 levels were ameliorated, while α-klotho levels increased.

**Conclusions:** Blood adjusted Ca, P, iPTH and α-klotho levels were correlated with Hb in CKD patients, correcting ROD is supposed to be therapeutic targets for anemia.

## Introduction

Renal anemia is a common complication in patients with chronic kidney disease (CKD). The incidence of anemia in non-dialysis patients exceeds 50% ^[1, 2]^, which is associated with increased morbidity and mortality related to cardiovascular disease, as well as increased risk of hospitalization^[1]^. Traditional causes of renal anemia include relative erythropoietin (EPO) deficiency, iron deficiency, and nutritional deficiency^[3]^. Correcting anemia is an important therapeutic target to improve the survival rate and quality of life for patients with CKD.

Chronic kidney disease-mineral and bone disorder (CKD-MBD) is a common complication and important cause of death in patients with CKD, characterized by dysregulated mineral and bone metabolism, bone abnormalities, and vascular calcification^[4, 5]^. Bone biopsy is the gold standard in the assessment of metabolic bone diseases, but it is invasive. Thus, bone related biomarkers have been clinically used to determine the bone turnover status in CKD and hemodialysis patients^[6]^. As the common manifestation of CKD-MBD, secondary hyperparathyroidism (SHPT) contributes to increased bone turnover, risk of fractures, vascular calcifications, cardiovascular and all-cause mortality^[7]^. Parathyroidectomy (PTX) is effective for treating severe SHPT patients^[8]^.

We have demonstrated that stage 5 CKD patients with severe SHPT had more significant anemia, which was significantly improved after parathyroidectomy (PTX)^[9]^. Cross-sectional studies suggest that there may be a link between biomarkers of mineral metabolism and hemoglobin (Hb)^[10-12]^. However, the correlations between systemic bone biomarkers and anemia in CKD patients remain unclear.

Serum calcium (Ca) and phosphorus (P) are related with bone metabolism. Alkaline phosphatase (ALP) plays an important role in skeletal mineralization and is the most widely recognized biochemical marker for osteoblast activity^[13]^. Intact parathyroid hormone (iPTH) is a major mediator of bone remodeling and an essential regulator of Ca and P homeostasis^[14]^. Fibroblast growth factor 23 (FGF23), a hormone secreted by osteocytes and osteoblasts, is a potent regulator of vitamin D metabolism and phosphate homeostasis^[11]^. Finally, α-klotho is a membrane protein that is highly expressed in the kidney, especially in the distal tubular epithelial cells^[15]^. Patients with advanced CKD have a significant reduction in α-klotho level and progressively lose the ability to prevent phosphate retention^[16]^.

The aim of this study is to investigate the relationships between blood bone metabolic biomarkers and anemia in CKD patients and analyze the effects of PTX on the above indices. This research will provide new insights for the pathogenesis and potential therapeutic targets for anemia in CKD patients.

## Materials and methods

### Study population

In this study, 239 CKD patients were enrolled from October 2011 to October 2018 from the Department of Nephrology, the First Affiliated Hospital of Nanjing Medical University. Among them 46 of all had severe SHPT and received total PTX with forearm auto-transplantation. A total of 28 PTX patients were followed up with a median interval of 6.88 months to investigate the effects of PTX on the blood bone metabolism indices and anemia. Healthy volunteers (n = 100) matched by age and sex were enrolled with the same exclusion criteria.

All of patients and controls participating in the study gave written informed consent. The study protocol was approved by the Research Ethics Committee of the First Affiliated Hospital of Nanjing Medical University, Nanjing, China (2015-SR-146).

### Inclusion criteria

The inclusion criteria of the patients included: (1) ages between 18-75 years old; (2) signed the written consent; (3) patients accord with the diagnostic criteria of CKD which are referenced from “KDIGO 2017 Clinical Practice Guideline Update for the Diagnosis, Evaluation, Prevention, and Treatment of Chronic Kidney Disease-Mineral and Bone Disorder (CKD-MBD)^[17]^ “.

### Exclusion criteria

The patients exhibited history of bleeding or blood transfusion, history of malignancy, serious infections, iron strengthening, lack of folic acid and vitamin B12, severe heart failure or active systemic disease 3 months prior to the observation and during the follow-up.

### PTX indications

(1) Persistent serum iPTH >800 pg/mL; (2) hypercalcemia and/or hyperphosphatemia that could not be controlled by medical therapy; (3) obvious clinical manifestations such as bone pain, pruritus, ectopic calcification or fracture; (4) at least one enlarged parathyroid gland discovered by ultrasound or a radiopharmaceutical technetium-99m-methoxyisobutylisonitrile (99mTc-MIBI) scan. Successful PTX is defined as blood iPTH levels<50 pg/mL within one week after surgery. Our previous study confirmed the mean percentage reduction of serum iPTH at 20 minutes after PTX (io-iPTH20%)>88.9% and the fourth day after PTX (D4-iPTH) ≤ 147.4 pg/mL could predict successful PTX^[18]^.

### Main observation indicators

During enrollment, we collected basic medical information about the CKD patients, such as demographics, dialysis mode, medical history, and causes of CKD. In addition, antihypertensive drugs for the patients were documented, including angiotensin converting enzyme inhibitor (ACEI), angiotensin receptor blocker (ARB), β-adrenergic receptor blocker, and calcium channel blockers (CCB). The personal information of healthy individuals was obtained through questionnaires.

Fasting venous blood in the morning was examined for routine blood tests, blood biochemistry, and related bone metabolism indices, including iPTH, FGF23 and α-klotho. The parameters were detected before and after PTX. Routine blood tests were performed using an LH-750 Hematology Analyzer (Beckman Coulter, Inc., Fullerton, CA). Biochemical indices were measured with an automatic biochemical analyzer (AU5400, Olympus Corporation, Japan). Serum iPTH was measured using a UniCel DxI 800 Access Immunoassay System (Beckman Coulter, Inc.). The related bone biomarkers were measured with human ELISA kits: plasma cFGF23 (Immutopics, San Clemente, CA); human soluble *α*-klotho (Immuno-Biologic Laboratories Co., Ltd. Japan).

Estimated glomerular filtration rate (eGFR) was calculated using the Modification of Diet in Renal Disease (MDRD) formula^[19]^. Serum calcium levels were corrected for serum albumin: adjusted calcium (mmol/l) = total calcium (mmol/L) + 0.02 × (40 − albumin [g/L]). Anemia was defined based on KDIGO clinical practice guideline for anemia in CKD when Hb <13.0 g/dL in men and <12.0 g/dL in women^[3]^. In this study, the degree of anemia was classified as mild, moderate, or severe based on the tertiles of Hb level of the anemic CKD patients, with cutoff values for the tertiles of 83g/L and 102g/L.

### Statistical analysis

The categorical variables were presented as numbers (proportions), and the continuous variables were presented as means± standard deviations (SD) or medians (interquartile range). The variables of the skewness distribution were transformed using the natural logarithm. Comparisons of normally distributed continuous variables were made with independent sample t tests or one-way ANOVA for continuous variables. A Mann-Whitney U test or Kruskal-Wallis test was used for skewed continuous variables and a chi-square test or exact probability test was performed for the categorical variables. Pearson correlation coefficient was used to analyze the correlation between various variables. To adjust for confounding factors, a single-variable regression analysis was first conducted to select the statistically significant variables (p<0.2) as independent variables. Subsequently, stepwise multivariate linear regression analysis was used to determine the factors independently associated with hemoglobin levels. Ordered logistic regression was performed to explore the effects of factors influencing the severity of anemia among CKD patients. A paired sampled t-test was used to assess the differences between the values recorded before and after the PTX. *P*<0.05 was considered statistically significant. All the statistical analyses were performed using the statistical package for the social sciences (SPSS), version 26.0 (SPSS Inc., Chicago, IL, USA).

## Results

### Characteristics of the study population

A total of 239 CKD patients and 100 healthy controls, matched for age and sex, were enrolled. The mean age and eGFR in 239 CKD patients were 49.25±13.22 years old and 25.74±35.99 ml/min/1.73 m^2^, including 130 males and 109 females. The most common cause of CKD was chronic glomerulonephritis (77.41%). Among the CKD patients, 58.58% did not receive dialysis, 32.22 % underwent hemodialysis, 9.21% had peritoneal dialysis and the dialysis vintage was 60.00(12.00-84.00) months. Of 99 dialysis patients, hemodialysis was the main dialysis mode. In our research, 30.54% patients used ACEI/ARB, 30.96% patients used active Vitamin D sterols, 52.72% patients took calcium channel blockers (CCB), and only 3.35% patients were treated with calcimimetics.

Compared with controls, the CKD patients had lower Hb, hematocrit (Hct), albumin (Alb), and higher triglycerides (TG), systolic blood pressure (SBP), and diastolic blood pressure (DBP). Anemia was observed in most CKD patients (84.10%). The baseline Hb level was 144.40±15.52 g/L in the controls and 98.76±25.62 g/L in the CKD patients.

Meanwhile, mineral and bone metabolism disorders were observed in CKD patients, with higher P, ALP, iPTH and FGF23 levels compared to controls. In addition, circulating Ca and α-klotho levels were significantly lower than controls. In order to eliminate pseudo-hypercalcemia, Serum calcium concentration was adjusted for serum albumin. The value of the albumin-adjusted calcium was 2.20±0.11 mmol/L in the controls and obviously increased in the CKD patients at 2.31±0.27mmol/L. The detailed comparisons of the control and CKD patients please see Table 1.

**Table 1.** Demographic data and laboratory values of controls and CKD patients.

| Variables | Controls<br>(n = 100) | CKD patients<br>(n = 239) | *p*<br>Value<sup>a</sup> | CKD patients |  |  |  | *p* Value<sup>b</sup> |
| --- | --- | --- | --- | --- | --- | --- | --- | --- |
|  |  |  |  | No anemia<br>(n = 38) | Mild anemia<br>(n = 70) | Moderate<br>anemia<br>(n=68) | Severe<br>anemia<br>(n=63) |  |
| Demographics |  |  |  |  |  |  |  |  |
| Age (years) | 48.97±13.54 | 49.25±13.22 | 0.859 | 45.24±13.04 | 51.07±12.31 | 49.91±12.90 | 48.94±14.39 | 0.169 |
| Male/female | 51/49 | 130/109 | 0.568 | 22/16 | 41/29 | 34/34 | 33/30 | 0.726 |
| Body mass index<br>(kg/m2) | 23.56±2.75 | 23.19±3.63 | 0.314 | 24.60±3.79 | 23.84±3.60 | 22.47±3.55 | 22.41±3.32 | **0.003** |
| Systolic pressure<br>(mmHg) | 122.63±16.36 | 146.00±25.92 | <0.001 | 132.24±16.92 | 144.96±25.16 | 149.87±22.44 | 153.51±30.99 | **<0.001** |
| Diastolic pressure<br>(mmHg) | 77.88±10.91 | 86.83±13.94 | <0.001 | 85.71±12.24 | 85.79 ±14.81 | 86.85±11.73 | 88.65±16.09 | 0.637 |
| Cause of ESRD, n (%) |  |  |  |  |  |  |  |  |
| Glomerulonephritis | / | 185(77.41) | / | 33(86.84) | 51(72.86) | 52(76.47) | 49(77.78) | 0.423 |
| Diabetic nephropathy | / | 24(10.04) | / | 2(5.26) | 9(12.86) | 7(10.29) | 6(9.52) | 0.629 |
| Hypertensive<br>nephropathy | / | 9(3.77) | / | 1(2.63) | 3(4.29) | 3(4.41) | 2(3.17) | 0.967 |
| Polycystic kidney<br>disease | / | 9(3.77) | / | 0(0) | 2(2.86) | 3(4.41) | 4(6.35) | / |
| other | / | 12(5.02) | / | 2(5.26) | 5(7.14) | 3(4.41) | 2(3.17) | 0.419 |
| Dialysis mode, n (%) |  |  |  |  |  |  |  |  |
| Non-dialysis | / | 140(58.58) | / | 34(89.47) | 45(64.29) | 29(42.65) | 32(50.79) | **<0.001** |
| Hemodialysis | / | 77(32.22) | / | 4(10.53) | 19(27.14) | 32(47.06) | 34(47.06) | **0.001** |
| Peritoneal dialysis | / | 22(9.21) | / | 0(0) | 6(8.57) | 7(10.29) | 9(14.29) | **0.027** |
| Dialysis vintage(m) | / | 60.00(12.0-84.0) | / | 102.00(30.00-168.00) | 60.00(17.00-108.50) | 60.00(24.00-96.00) | 36.00(5.00-72.00) | 0.066 |
| **Medication history, n (%)** |  |  |  |  |  |  |  |  |
| CCB | / | 126(52.72) | / | 13(34.21) | 34(48.57) | 42(61.76) | 37(58.73) | **0.031** |
| ACEI/ARB | / | 73(30.54) | / | 16(42.11) | 22(31.43) | 22(32.35) | 13(20.63) | 0.142 |
| β-receptor blocker | / | 53(22.18) | / | 5(13.16) | 16(22.86) | 17(25.00) | 15(23.81) | 0.528 |
| Active vitamin D sterols | / | 74(30.96) | / | 4(10.53) | 18(25.71) | 33(48.53) | 19(30.16) | **<0.001** |
| Calcimimetics | / | 8(3.35) | / | 1(2.63) | 4(5.71) | 3(4.41) | 0(0.00) | 0.264 |
| **Laboratory values** |  |  |  |  |  |  |  |  |
| Hemoglobin (g/L) | 144.40±15.52 | 98.76±25.62 | <0.001 | 139.39±13.63 | 110.86±5.99 | 92.71±5.96 | 67.38±10.56 | **<0.001** |
| Hematocrit (%) | 43.26±4.24 | 30.19±7.66 | <0.001 | 41.93±3.98 | 33.96±2.12 | 28.41±2.18 | 20.83±3.56 | **<0.001** |
| Glucose (mmol/L) | 5.41±0.77 | 5.28±2.33 | 0.442 | 5.58±2.82 | 5.64±2.92 | 4.87±1.30 | 5.14±2.10 | 0.202 |
| Albumin (g/L) | 47.76±2.87 | 35.74±6.63 | <0.001 | 37.14±7.59 | 36.36±6.92 | 34.57±7.13 | 35.50±4.78 | 0.212 |
| Creatinine (μmol/L) | 71.75±15.61 | 625.65±462.33 | <0.001 | 206.51±257.33 | 471.02±400.79 | 705.20±375.35 | 964.40±437.92 | **<0.001** |
| Urea (mmol/L) | 5.56±1.42 | 20.16±11.90 | <0.001 | 9.14±7.15 | 15.68±8.07 | 21.39±8.55 | 30.45±12.47 | **<0.001** |
| Total cholesterol (mmol/L) | 5.01±0.83 | 4.98±1.89 | 0.816 | 5.92±2.90 | 5.33±1.77 | 4.76±1.51 | 4.27±1.25 | **<0.001** |
| Triglyceride (mmol/L) | 1.44±1.45 | 1.92±1.33 | 0.003 | 2.02±1.02 | 2.22±1.49 | 1.90±1.39 | 1.53±1.16 | **0.024** |
| eGFR (ml/min/1.73 m<sup>2</sup>) | 97.78±18.26 | 25.74±35.99 | <0.001 | 65.46±45.13 | 34.69±40.53 | 12.48±15.88 | 6.16±4.11 | **<0.001** |
| Calcium (mmol/L) | 2.36±0.11 | 2.23±0.28 | <0.001 | 2.28±0.17 | 2.25±0.30 | 2.26±0.27 | 2.13±0.32 | **0.021** |
| Adjusted Ca (mmol/L) | 2.20±0.11 | 2.31±0.27 | <0.001 | 2.34±0.15 | 2.32±0.26 | 2.37±0.23 | 2.22±0.31 | **0.013** |
| Phosphorus (mmol/L) | 1.17±0.15 | 1.72±0.60 | <0.001 | 1.26±0.38 | 1.55±0.48 | 1.86±0.54 | 2.04±0.66 | **<0.001** |
| lnALP | 4.31±0.26 | 4.63±0.81 | <0.001 | 4.40±0.69 | 4.56±0.72 | 4.81±0.94 | 4.64±0.80 | 0.076 |
| lniPTH | 3.58±0.39 | 5.18±1.61 | <0.001 | 4.04±1.36 | 4.59±1.58 | 5.72±1.56 | 5.94±1.14 | **<0.001** |
| lnFGF23 | 4.15±0.31 | 7.29±2.36 | <0.001 | 5.29±1.72 | 6.79±2.29 | 8.20±2.40 | 8.07±1.82 | **<0.001** |
| lnKlotho | 6.50±0.27 | 5.82±0.49 | <0.001 | 6.05±0.44 | 5.78±0.50 | 5.80±0.52 | 5.74±0.43 | **0.012** |

Data are presented as means ± SD or median (interquartile range) for continuous variables and as n (%) for categorical variables.
Abbreviations: CKD, chronic kidney disease; BMI, body mass index; SBP, systolic blood pressure; DBP, diastolic blood pressure; CCB, calcium channel blocker; ACEI/ARB, angiotensin converting enzyme inhibitors/angiotensin receptor blocker; Ca, calcium; TC, total cholesterol; TG, triglyceride; eGFR, estimated glomerular filtration rate; ALP, alkaline phosphatase; iPTH, intact parathyroid hormone; FGF23, fibroblast growth factor 23.
\*Natural log-transformed. <sup>a</sup>Controls vs. CKD patients. <sup>b</sup>Analysis of variance of 4 groups in CKD patients.

#### The correlations between blood bone biomarkers and anemia in CKD patients

The CKD patients were divided into four subgroups according to the Hb levels: no anemia (male: Hb≥130g/L, female: Hb≥120g/L, n=38), mild anemia (male: 102≤Hb<130g/L, female: 102≤Hb<120g/L, n=70), moderate anemia (83≤Hb<102g/L, n=68), severe anemia (30≤Hb<83g/L, n=63). The baseline Hb levels in the no anemia, mild, moderate, and severe anemia subgroups were 139.39±13.63g/L, 110.86±5.99g/L, 92.71±5.96g/L, and 67.38±10.56g/L, respectively. Patients with more severe anemia exhibited higher levels of P, lnALP, lniPTH and lnFGF23. Contrarily, serum adjusted Ca and α-klotho levels showed a decreasing trend. Lower BMI and albumin levels, higher SBP were observed in CKD patients with more severe anemia (Table 1, Figure 1).

**Figure 1.**
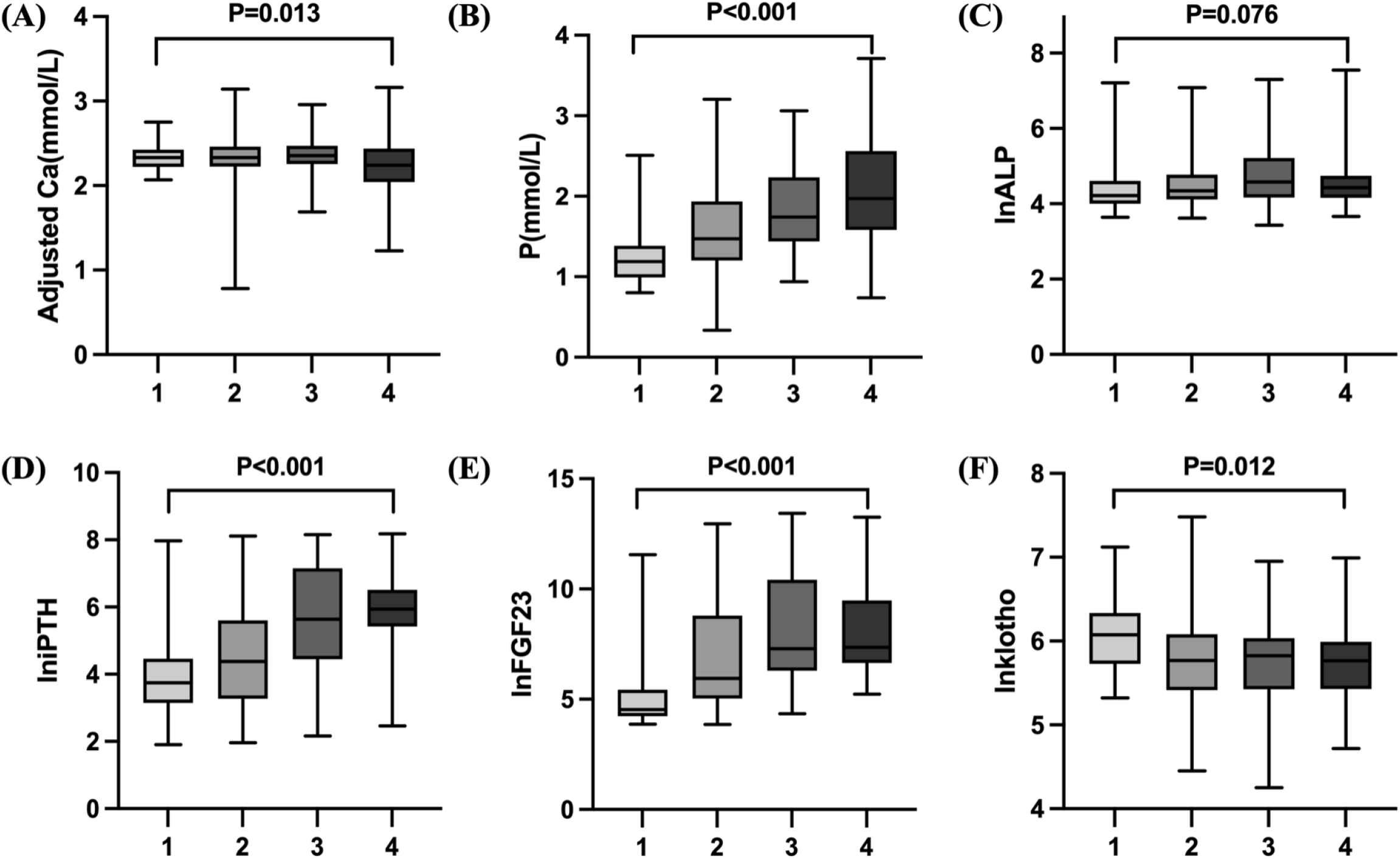
Baseline blood bone metabolism indices in different subgroups of CKD patients. The CKD patients were divided into three subgroups according to the hemoglobin levels: 1: no anemia(male: Hb⩾130g/L;female: Hb⩾120g/L, n=38), 2: mild anemia(male: 102⩽Hb<130g/L;female: 102⩽Hb<120g/L, n=70), 3: moderate anemia(83⩽Hb<102g/L, n=68), 4: severe anemia(30⩽Hb<83g/L, n=63). Abbreviations: Ca, calcium; P, phosphorus; ALP, alkaline phosphatase; iPTH, intact parathyroid hormone; FGF23, fibroblast growth factor 23.

Correlations between blood bone metabolism indexes and Hb in 239 CKD patients are illustrated in Figure 2. At baseline, Hb levels had significant negative correlations with lniPTH, lnFGF23 and P levels (P<0.001). Meanwhile, positive correlation between ln(α-klotho) and Hb was revealed (r=0.171, p=0.01). Hb was shown to be correlated with serum adjusted Ca levels (r=0.130, p=0.044). Furthermore, sex, BMI, SBP, DBP, TC and TG were correlated with Hb (data not shown).

**Figure 2.**
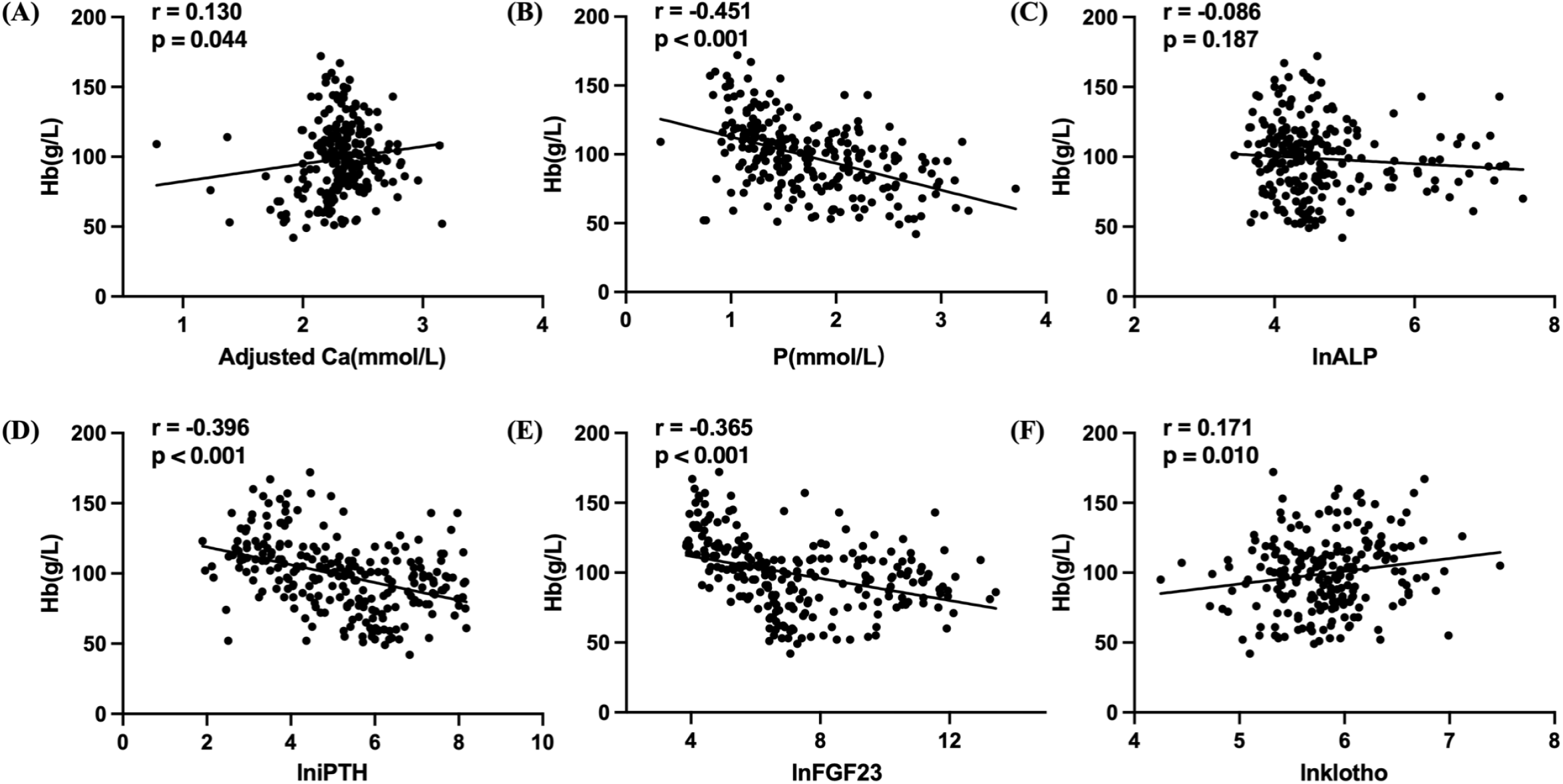
Correlations between blood bone metabolism indices and Hb levels in CKD patients. Abbreviations: Hb, hemoglobin; Ca, calcium; P, phosphorus; ALP, alkaline phosphatase; iPTH, intact parathyroid hormone; FGF23, fibroblast growth factor 23.

#### The independent influencing factors of anemia severity in CKD patients

To eliminate the influence of potential confounding factors, a multivariable stepwise linear regression analysis with Hb as dependent variable was established. In CKD patients, the Hb levels were inversely correlated with blood P levels (β = -0.264, P <0.001) and lniPTH (β = -0.157, P=0.026). Serum adjusted Ca and ln(α-klotho) were positively related with Hb levels. There were no correlations between circulating FGF23 and Hb levels. Sex, BMI, SBP, Alb and TC were independently correlated with Hb in CKD patients (Table 2).

**Table 2.** Influencing factors of Hb levels in CKD patients analyzed by multiple stepwise regression.

| Independent Variable | Unstandardized Coefficient | Standardized Coefficient | 95% CI |  | *p* Value |
| --- | --- | --- | --- | --- | --- |
|  |  |  | Lower | Upper |  |
| **Female sex** | -11.690 | -0.224 | -17.117 | -6.263 | <0.001 |
| **BMI** | 0.955 | 0.130 | 0.200 | 1.710 | 0.013 |
| **SBP** | -0.170 | -0.170 | -0.275 | -0.064 | 0.002 |
| **Alb** | 1.400 | 0.355 | 0.957 | 1.844 | <0.001 |
| **TC** | 3.990 | 0.296 | 2.423 | 5.558 | <0.001 |
| **Adjusted ca** | 12.903 | 0.134 | 3.019 | 22.786 | 0.011 |
| **P** | -11.505 | -0.264 | -17.216 | -5.794 | <0.001 |
| **lniPTH** | -2.574 | -0.157 | -4.840 | -0.307 | 0.026 |
| **lnklotho** | 5.622 | 0.105 | 0.151 | 11.093 | 0.044 |

Multiple stepwise regression model with adjustment for age, sex, BMI, SBP, DBP, and Glu, Alb, TC, TG, Ca, P, lniPTH, lnFGF23, lnklotho and CCB, ACEI/ARB, $\beta$ -receptor blocker, calcimimetics and active Vitamin D sterols.
Abbreviations: Hb, hemoglobin; CKD, chronic kidney disease; BMI, body mass index; SBP, systolic blood pressure; Alb, albumin; TC, total cholesterol; Ca, calcium; P, phosphorus; iPTH, intact parathyroid hormone; CI, confidence interval.

These findings were further strengthened in a multivariate ordered logistic regression. The anemia level was analyzed based on mild, moderate and severe anemia subgroups. After rigorous adjustment of confounders, including age, sex, BMI, SBP, DBP, medication history, and biochemical indicators, significant factors identified for the severity of anemia in CKD patients included serum adjusted Ca [odds ratio (OR) = 0.216, 95% confidence interval (CI) = 0.068-0.684; *P* = 0.009], phosphorus (OR = 3.019, 95% CI = 1.621-5.624; *P* = 0.001) and lniPTH (OR = 1.366, 95% CI = 1.005-1.857; *P* = 0.046). Similarly, sex, SBP, Alb and TC were all independent factors for severity of anemia (Figure 3).

**Figure 3.**
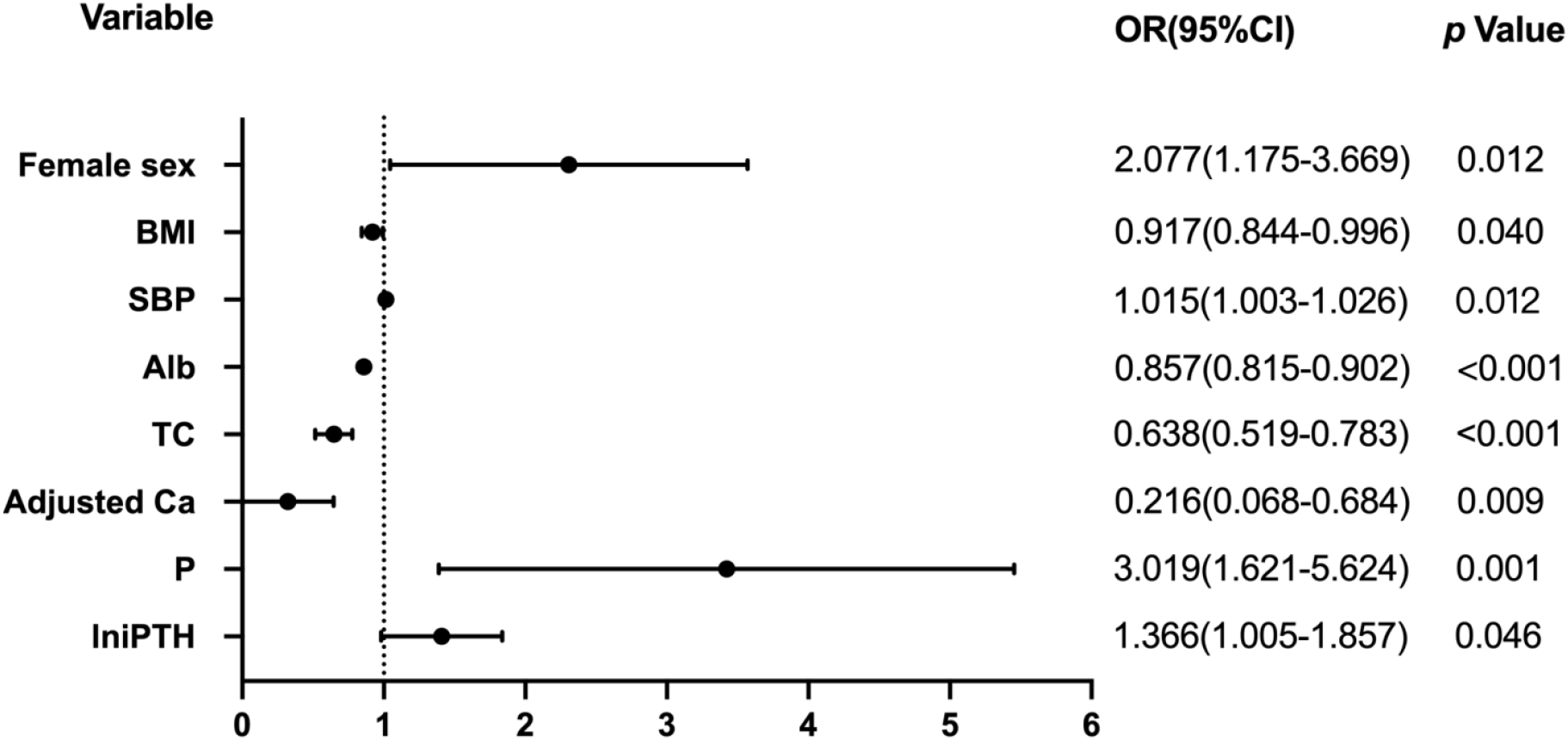
Multiple ordered logistic regression analysis of factors associated with anemia in CKD patients. Abbreviations: BMI, body mass index; SBP, systolic blood pressure; Alb, albumin; TC, total cholesterol; Ca, calcium; P, phosphorus; iPTH, intact parathyroid hormone; OR, odds ratio; CI, confidence interval.

#### Effects of PTX on laboratory indicators and bone metabolism indices in patients with severe SHPT

CKD patients were categorized into stages 1 to 5 based on eGFR and the numbers of patients in each stage was 17, 21, 21, 26 and 154, respectively. Among the stage 5 CKD subgroup, 46 patients with severe SHPT underwent PTX, whom were younger, and hemodialysis was the main dialysis mode compared to the patients without PTX.

A total of 28 PTX patients were followed up (median time: 6.88 months). Although the duration of follow-up differed in PTX patients, blood bone metabolism disorders were alleviated, and no time trend were observed (data not shown). After PTX, anemia, hypoalbuminemia and bone metabolism disorders were improved significantly. Compared with baseline data, the levels of adjusted Ca, P, iPTH and FGF23 decreased, while the levels of α-klotho increased in the patients after PTX (Figure 4). We further investigated the correlations between the longitudinal improvements of blood bone biomarkers and Hb levels in PTX patients, however, no correlation was found (Table 3).

**Figure 4.**
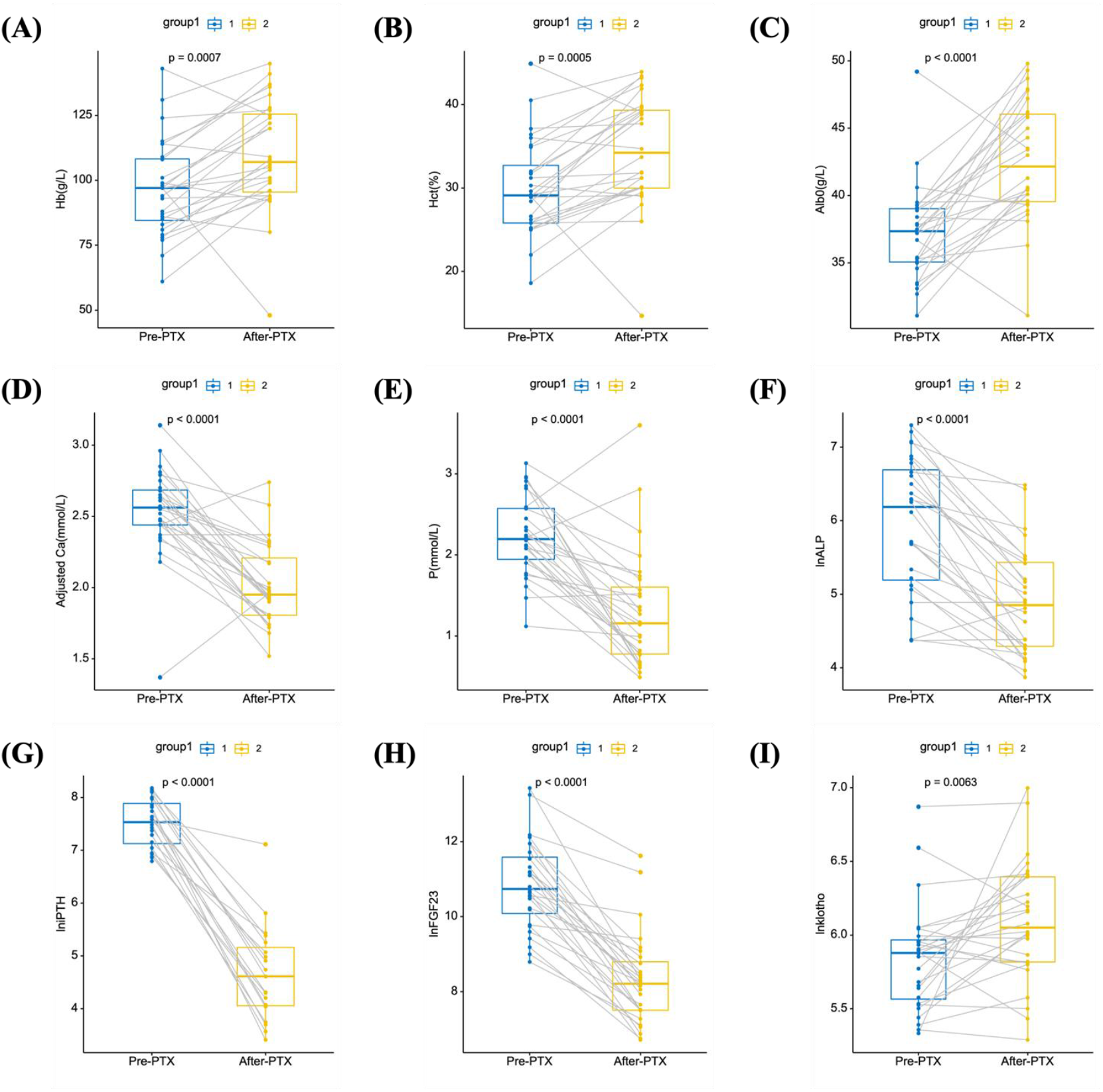
Comparisons of laboratory indices in severe SHPT patients undergoing PTX. Abbreviations: Hb, hemoglobin; Hct, hematocrit; Alb, albumin; Ca, calcium; P, phosphorus; ALP, alkaline phosphatase; iPTH, intact parathyroid hormone; FGF23, fibroblast growth factor 23.

**Table 3.**
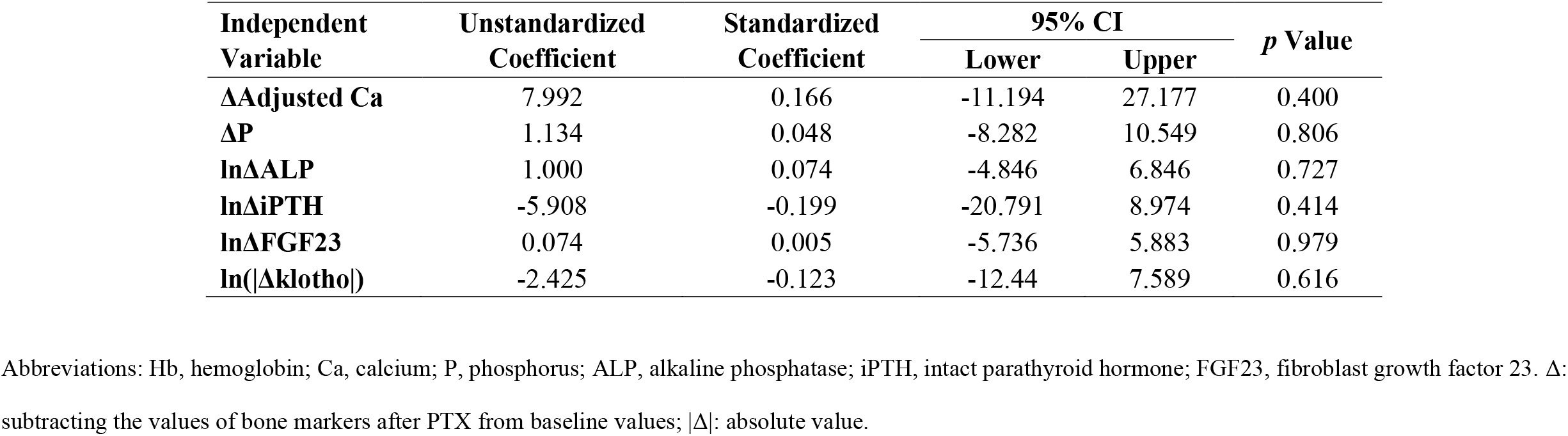
Correlations between the changes of bone metabolism indices and Hb levels in PTX patients before and after operation.

## Discussion

The present investigation was designed to analyze the relationships between blood bone biomarkers and anemia in CKD patients and to assess how PTX affected these variables in the severe SHPT subgroup. Hypoalbuminemia is recognized as a strong predictor of mortality in the CKD population^[20]^. In our research, serum albumin level was an independent factor of Hb levels in CKD patients, and Hb was also affected by sex, BMI, blood pressure and blood lipids^[21, 22]^.

We proved that both serum calcium and phosphorus were independently associated with Hb levels in CKD patients. Mauro et al^[22]^ reported that the decrease of serum Ca and the increase of serum phosphorus levels were closely related to anemia in patients with advanced non-dialysis CKD. Calcium was required for the *in vitro* proliferation and differentiation of erythroid progenitor cells induced by erythropoietin^[23]^. As erythropoietin activated pathway, store-operated calcium channel (SOC) related genetic polymorphisms are proved to be correlated with the risk of EPO resistance in dialysis patients^[24]^.

The correlations between hyperphosphatemia and anemia are also reported^[12, 25]^. Erythropoietin deficiency, inflammation and oxidative stress have been implicated as potential factors linking hyperphosphatemia and anemia. Hyperphosphatemia may increase the synthesis of polyamines, which can act as toxins inhibiting erythropoiesis^[25]^. Hyperphosphatemia in CKD can decrease vitamin D synthesis, resulting in hypocalcemia and elevated PTH levels, which further inhibit erythropoiesis^[26]^. FGF-23 and α-klotho may play important roles in the mechanism underlying the association between hyperphosphatemia and anemia^[12, 27]^. High serum phosphorus can increase FGF-23 levels, which in turn suppresses the activity of α-klotho and aggravates vitamin D deficiency^[27, 28]^. Klotho deficiency increases P reabsorption in the kidneys, which results in hyperphosphatemia^[29]^. Low levels of α-klotho and vitamin D are implicated with an increased risk of anemia^[30, 31]^.

A positive association between serum ALP levels and erythropoiesis stimulating agent (ESA)-resistant anemia has been demonstrated in a study involving 38,328 end-stage kidney disease (ESKD) patients receiving hemodialysis^[13]^. Serum ALP levels are associated with severity of resistance to ESA in CKD patients with no identifiable causes^[32]^. Previous studies have demonstrated that ALP is associated with mortality in ESKD patients receiving dialysis^[33]^ and pre-dialysis patients with CKD^[34, 35]^. To the best of our knowledge, the relationship between serum ALP levels and Hb in CKD patients has not been investigated. However, in our study, no significant correlation was identified between ALP and Hb levels in CKD populations, which may be related to the less number of cases and lack of information on the use of ESA.

It has been recognized that increased iPTH can inhibit EPO synthesis, shorten erythrocyte survival time, and lead to bone marrow fibrosis, thus reducing hematopoietic function^[36]^. However, the molecular mechanism of high PTH inhibiting the synthesis of endogenous EPO and myeloid progenitor cells remains unclear. In our study, baseline iPTH levels were independently associated with Hb levels and severity of anemia in CKD patients. SHPT is a common complication of advanced CKD and a recognized cause of ESA resistance. Kamalas et al. ^[37]^ conducted a retrospective study on 43 anemic CKD patients who received ESA treatment and found that in patients with normal bone marrow examination, the higher iPTH levels had lower Hb levels and needed higher doses of ESA, suggesting that the low reactivity of ESA in CKD patients was related to high levels of iPTH.

We proved that blood FGF23 levels were negatively correlated with the Hb levels of CKD patients. A recent report from the Chronic Renal Insufficiency Cohort (CRIC) study showed that increased FGF23 levels were significantly correlated with decreased Hb and overall rates of anemia^[38]^. Moreover, elevated serum FGF23 levels in CKD patients are negatively correlated with transferrin saturation, serum iron levels, and erythropoiesis^[39, 40]^. Coe et al ^[41]^ observed a significant increase in the number of hematopoietic stem cells (HSCs) and erythroid progenitor cells in bone marrow and peripheral blood in FGF23 deficient mice. Elevated serum FGF23 levels can stimulate the synthesis and secretion of various pro-inflammatory cytokines (such as IL-6 and IL-1β). Cytokines up-regulate the production of ferritin in the liver, which in turn promotes the high level of serum ferritin, leading to functional iron deficiency^[42]^.

The anti-aging gene α-klotho is an important marker of CKD, and its corresponding protein is mainly expressed in the kidney^[15]^. Previous research has found that Hct was significantly decreased in klotho gene mutant (*KL*^-/-^) mice, indicating that klotho deficiency caused anemia was mediated by suppressing the expression of *S*-formylglutathione hydrolase (FGH)^[43]^. Klotho gene deficiency in kidneys leads to decreased plasma klotho levels, which suppresses FGH expression in red blood cells (RBCs). FGH is a key enzyme in the generation of the major cellular antioxidant glutathione (GSH), which is involved in the maintenance of normal homeostasis of RBCs and kidneys. A decrease in the GSH levels would damage RBCs and lead to anemia^[43-45]^. The study also observed glomerular collapse and interstitial fibrosis in *KL*^-/-^ mice, indicating that haplodeficiency of klotho gene (*K*) impaired kidney function, which in turn contributed to klotho deficiency-related anemia. In addition, full-length α-klotho is reported to have an inhibitory effect on oxidative stress, which may help to alleviate anemia^[25, 29]^. Sangeetha et al^[46]^ demonstrated that loss of klotho disrupted erythropoiesis and HSCs development. These hematopoietic changes of bone marrow could be caused by the dramatic reduction in osteoblast and osteoclast numbers and bone mineral density in *Klotho*^−/−^ bones, which may be responsible for the inhibition of hypoxia inducible factor (HIF) and EPO in bone.

Klotho deficiency is a characteristic feature of CKD, however, the role of α-klotho in renal anemia remains unclear. Our study demonstrated that blood α-klotho levels were positively associated with Hb levels in CKD patients. The treatment of anemia with EPO has been demonstrated to enhance renal and extrarenal production of α-klotho in CKD patients^[30]^. However, several studies held the opposite opinion. Yang Xu et al^[31]^ suggested that there may be a negative feedback between EPO and α-klotho such that increasing α-klotho suppresses the production of EPO in CKD, thus low expression of α-klotho may be a compensatory mechanism to attenuate the effects of anemia. Another study found that the expression of HIF-1 α and HIF-2 α is significantly up-regulated in α-klotho-/- bone tissue, resulting in local overexpression of EPO^[47]^. Although the specific mechanism of α-klotho in renal anemia remains unclear, it is certain that α-klotho is a therapeutic target for renal anemia. The relationship between α-klotho and renal anemia is also affected by iron metabolism and vitamin D, which requires more research to draw the conclusions^[31]^.

Clinical studies have confirmed the importance of treating SHPT in improving renal anemia in hemodialysis patients^[9, 48]^. Zingraff et al^[49]^ have demonstrated that successful PTX can significantly improve anemia in SHPT patients, consistent with the conclusions in our previous reports^[9, 50]^. In this study, PTX patients(n=28) were followed up over three months after the surgery (median time: 6.88 months). Elevated serum phosphorus, iPTH, ALP, FGF23 and decreased α-klotho levels were alleviated after PTX, however, we did not find the correlations between correction of bone metabolism disorders and improvement of anemia after PTX.

There are several limitations in this study. First, direct cause-and-effects relationships between the blood bone metabolic biomarkers and anemia in CKD patients could not be confirmed. Second, we did not collect the medication history of ESA and blood transfusion, iron metabolism related indexes in CKD patients. Third, bone histology analysis was not performed and less PTX patients were followed up in the study which might be related with an unmeasurable bias.

## Conclusions

Anemia and CKD-MBD are both common complications in CKD patients, which are strongly correlated with poor prognosis. Here we proved that in CKD patients, increased blood adjusted Ca, P, iPTH and decreased α-klotho levels were independently associated with reduced Hb levels. Moreover, adjusted Ca, P and iPTH were independent factors affecting the severity of renal anemia. Our data indicated that PTX could correct the anemia and bone metabolic disorders in patients with severe SHPT, but no correlation was found between the increased Hb levels and different corrected bone metabolism biomarkers. This study provides a reference for in-depth analysis of the pathogenesis and therapeutic targets for renal anemia, from the perspective of CKD-MBD.

## Data Availability

All data produced in the present study are available upon reasonable request to the authors.

## Acknowledgement

The authors sincerely thank the patients and their families for participation in our study. The authors thank all medical staffs and students involved in the clinical management and analysis of samples for the patients.

## References

[1] Toft G, Heide-Jørgensen U, Van Haalen H, et al. Anemia and clinical outcomes in patients with non-dialysis dependent or dialysis dependent severe chronic kidney disease: a Danish population-based study [J]. J Nephrol, 2020, 33(1): 147–56.

[2] Vestergaard S V, Heide-Jørgensen U, Van Haalen H, et al. Risk of Anemia in Patients with Newly Identified Chronic Kidney Disease - A Population-Based Cohort Study [J]. Clin Epidemiol, 2020, 12: 953–62.

[3] Kidney Disease: Improving Global Outcomes (KDIGO) Anemia Work Group. KDIGO Clinical Practice Guideline for Anemia in Chronic Kidney Disease. [J]. Kidney inter, Suppl 2012, 2012; 2: 279–335.

[4] Hruska K A, Sugatani T, Agapova O, et al. The chronic kidney disease - Mineral bone disorder (CKD-MBD): Advances in pathophysiology [J]. Bone, 2017, 100: 80–6.

[5] Ketteler M, Block G A, Evenepoel P, et al. Diagnosis, Evaluation, Prevention, and Treatment of Chronic Kidney Disease-Mineral and Bone Disorder: Synopsis of the Kidney Disease: Improving Global Outcomes 2017 Clinical Practice Guideline Update [J]. Ann Intern Med, 2018, 168(6): 422–30.

[6] Zheng C M, Wu C C, Lu C L, et al. Hypoalbuminemia differently affects the serum bone turnover markers in hemodialysis patients [J]. Int J Med Sci, 2019, 16(12): 1583–92.

[7] Wan J, Li W, Zhong Y. Parathyroidectomy decreases serum intact parathyroid hormone and calcium levels and prolongs overall survival in elderly hemodialysis patients with severe secondary hyperparathyroidism [J]. J Clin Lab Anal, 2019, 33(3): e22696.

[8] Sun X, Zhang X, Lu Y, et al. Risk factors for severe hypocalcemia after parathyroidectomy in dialysis patients with secondary hyperparathyroidism [J]. Sci Rep, 2018, 8(1): 7743.

[9] Chen H, Ren W, Gao Z, et al. Effects of parathyroidectomy on plasma PTH fragments and heart rate variability in stage 5 chronic kidney disease patients [J]. Ren Fail, 2021, 43(1): 890–9.

[10] Tsai M H, Leu J G, Fang Y W, et al. High Fibroblast Growth Factor 23 Levels Associated With Low Hemoglobin Levels in Patients With Chronic Kidney Disease Stages 3 and 4 [J]. Medicine (Baltimore), 2016, 95(11): e3049.

[11] Mehta R, Cai X, Hodakowski A, et al. Fibroblast Growth Factor 23 and Anemia in the Chronic Renal Insufficiency Cohort Study [J]. Clin J Am Soc Nephrol, 2017, 12(11): 1795–803.

[12] Tran L, Batech M, Rhee C M, et al. Serum phosphorus and association with anemia among a large diverse population with and without chronic kidney disease [J]. Nephrol Dial Transplant, 2016, 31(4): 636–45.

[13] Kalantar-Zadeh K, Lee G H, Miller J E, et al. Predictors of hyporesponsiveness to erythropoiesis-stimulating agents in hemodialysis patients [J]. Am J Kidney Dis, 2009, 53(5): 823–34.

[14] Tabibzadeh N, Karaboyas A, Robinson B M, et al. The risk of medically uncontrolled secondary hyperparathyroidism depends on parathyroid hormone levels at haemodialysis initiation [J]. Nephrol Dial Transplant, 2021, 36(1): 160–9.

[15] Thongprayoon C, Neyra J A, Hansrivijit P, et al. Serum Klotho in Living Kidney Donors and Kidney Transplant Recipients: A Meta-Analysis [J]. J Clin Med, 2020, 9(6).

[16] Maique J, Flores B, Shi M, et al. High Phosphate Induces and Klotho Attenuates Kidney Epithelial Senescence and Fibrosis [J]. Front Pharmacol, 2020, 11: 1273.

[17] KDIGO 2017 Clinical Practice Guideline Update for the Diagnosis, Evaluation, Prevention, and Treatment of Chronic Kidney Disease-Mineral and Bone Disorder (CKD-MBD) [J]. Kidney Int Suppl (2011), 2017, 7(1): 1–59.

[18] Zhang L, Xing C, Shen C, et al. Diagnostic Accuracy Study of Intraoperative and Perioperative Serum Intact PTH Level for Successful Parathyroidectomy in 501 Secondary Hyperparathyroidism Patients [J]. Sci Rep, 2016, 6: 26841.

[19] Levey A S, Bosch J P, Lewis J B, et al. A more accurate method to estimate glomerular filtration rate from serum creatinine: a new prediction equation. Modification of Diet in Renal Disease Study Group [J]. Ann Intern Med, 1999, 130(6): 461–70.

[20] Kovesdy C P, Kalantar-Zadeh K. Why is protein-energy wasting associated with mortality in chronic kidney disease? [J]. Semin Nephrol, 2009, 29(1): 3–14.

[21] Alemayehu M, Meskele M, Alemayehu B, et al. Prevalence and correlates of anemia among children aged 6-23 months in Wolaita Zone, Southern Ethiopia [J]. PLoS One, 2019, 14(3): e0206268.

[22] Boronat M, Santana Á, Bosch E, et al. Relationship between Anemia and Serum Concentrations of Calcium and Phosphorus in Advanced Non-Dialysis-Dependent Chronic Kidney Disease [J]. Nephron, 2017, 135(2): 97–104.

[23] Misiti J, Spivak J L. Erythropoiesis in vitro. Role of calcium [J]. J Clin Invest, 1979, 64(6): 1573–9.

[24] Kao C C, Wong H S, Wang Y J, et al. The role of genetic polymorphisms in STIM1 and ORAI1 for erythropoietin resistance in patients with renal failure [J]. Medicine (Baltimore), 2021, 100(17): e25243.

[25] Kovesdy C P, Mucsi I, Czira M E, et al. Association of serum phosphorus level with anemia in kidney transplant recipients [J]. Transplantation, 2011, 91(8): 875–82.

[26] Blaine J, Chonchol M, Levi M. Renal control of calcium, phosphate, and magnesium homeostasis [J]. Clin J Am Soc Nephrol, 2015, 10(7): 1257–72.

[27] Krajisnik T, Olauson H, Mirza M A, et al. Parathyroid Klotho and FGF-receptor 1 expression decline with renal function in hyperparathyroid patients with chronic kidney disease and kidney transplant recipients [J]. Kidney Int, 2010, 78(10): 1024–32.

[28] Razzaque M S. Interactions between FGF23 and vitamin D [J]. Endocr Connect, 2022, 11(10).

[29] Xu Y, Sun Z. Molecular basis of Klotho: from gene to function in aging [J]. Endocr Rev, 2015, 36(2): 174–93.

[30] Milovanov Y S, Mukhin N A, Kozlovskaya L V, et al. [Impact of anemia correction on the production of the circulating morphogenetic protein α-Klotho in patients with Stages 3B-4 chronic kidney disease: A new direction of cardionephroprotection] [J]. Ter Arkh, 2016, 88(6): 21–5.

[31] Xu Y, Peng H, Ke B. alpha-klotho and anemia in patients with chronic kidney disease patients: A new perspective [J]. Exp Ther Med, 2017, 14(6): 5691–5.

[32] Badve S V, Zhang L, Coombes J S, et al. Association between serum alkaline phosphatase and primary resistance to erythropoiesis stimulating agents in chronic kidney disease: a secondary analysis of the HERO trial [J]. Can J Kidney Health Dis, 2015, 2: 33.

[33] Rhee C M, Molnar M Z, Lau W L, et al. Comparative mortality-predictability using alkaline phosphatase and parathyroid hormone in patients on peritoneal dialysis and hemodialysis [J]. Perit Dial Int, 2014, 34(7): 732–48.

[34] Beige J, Wendt R, Girndt M, et al. Association of serum alkaline phosphatase with mortality in non-selected European patients with CKD5D: an observational, three-centre survival analysis [J]. BMJ Open, 2014, 4(2): e004275.

[35] Taliercio J J, Schold J D, Simon J F, et al. Prognostic importance of serum alkaline phosphatase in CKD stages 3-4 in a clinical population [J]. Am J Kidney Dis, 2013, 62(4): 703–10.

[36] Tanaka M, Komaba H, Fukagawa M. Emerging Association Between Parathyroid Hormone and Anemia in Hemodialysis Patients [J]. Ther Apher Dial, 2018, 22(3): 242–5.

[37] Amnuay K, Srisawat N, Wudhikarn K, et al. Factors associated with erythropoiesis-stimulating agent hyporesponsiveness anemia in chronic kidney disease patients [J]. Hematol Rep, 2019, 11(3): 8183.

[38] Mehta R, Cai X, Hodakowski A, et al. Fibroblast Growth Factor 23 and Anemia in the Chronic Renal Insufficiency Cohort Study [J]. Clin J Am Soc Nephrol, 2017, 12(11): 1795–803.

[39] Lewerin C, Ljunggren Ö, Nilsson-Ehle H, et al. Low serum iron is associated with high serum intact FGF23 in elderly men: The Swedish MrOS study [J]. Bone, 2017, 98: 1–8.

[40] Agoro R, Montagna A, Goetz R, et al. Inhibition of fibroblast growth factor 23 (FGF23) signaling rescues renal anemia [J]. Faseb j, 2018, 32(7): 3752–64.

[41] Coe L, Madathil S, Casu C, et al. FGF-23 is a negative regulator of prenatal and postnatal erythropoiesis [J]. The Journal of biological chemistry, 2014, 289(14): 9795–810.

[42] Czaya B, Faul C. The Role of Fibroblast Growth Factor 23 in Inflammation and Anemia [J]. Int J Mol Sci, 2019, 20(17).

[43] Xu Y, Sun Z. Regulation of S-formylglutathione hydrolase by the anti-aging gene klotho [J]. Oncotarget, 2017, 8(51): 88259–75.

[44] Ghashghaeinia M, Giustarini D, Koralkova P, et al. Pharmacological targeting of glucose-6-phosphate dehydrogenase in human erythrocytes by Bay 11-7082, parthenolide and dimethyl fumarate [J]. Sci Rep, 2016, 6: 28754.

[45] Ellison I, Richie J P, Jr. Mechanisms of glutathione disulfide efflux from erythrocytes [J]. Biochem Pharmacol, 2012, 83(1): 164–9.

[46] Vadakke Madathil S, Coe L, Casu C, et al. Klotho deficiency disrupts hematopoietic stem cell development and erythropoiesis [J]. The American journal of pathology, 2014, 184(3): 827–41.

[47] Hu M C, Shi M, Zhang J, et al. Renal Production, Uptake, and Handling of Circulating αKlotho [J]. J Am Soc Nephrol, 2016, 27(1): 79–90.

[48] Tanaka M, Yoshida K, Fukuma S, et al. Effects of Secondary Hyperparathyroidism Treatment on Improvement in Anemia: Results from the MBD-5D Study [J]. PLoS One, 2016, 11(10): e0164865.

[49] Zingraff J, Drüeke T, Marie P, et al. Anemia and secondary hyperparathyroidism [J]. Arch Intern Med, 1978, 138(11): 1650–2.

[50] Jiang Y, Zhang J, Yuan Y, et al. Association of Increased Serum Leptin with Ameliorated Anemia and Malnutrition in Stage 5 Chronic Kidney Disease Patients after Parathyroidectomy [J]. Sci Rep, 2016, 6: 27918.

